# Exploring Risk Factors for Comorbid Depression in Osteoarthritis: A Scoping Review Protocol

**DOI:** 10.1101/2024.08.26.24312577

**Authors:** Kathrin Bogensberger, Dagmar Schaffler-Schaden, Eva Perl, Markus Ritter, Wolfgang Hitzl, Bibiane Steinecker-Frohnwieser, Antje van der Zee-Neuen

**Affiliations:** Ludwig Boltzmann Institute for Arthritis and Rehabilitation, Vienna, Austria; Center for Physiology, Pathophysiology and Biophysics, Institute of Physiology and Pathophysiology, Paracelsus Medical University, Salzburg, Austria; Gastein Research Institute, Paracelsus Medical University, Salzburg, Austria; Institute of General Practice, Family Medicine and Preventive Medicine, Paracelsus Medical University Salzburg; Department of Geriatric Medicine, Christian-Doppler Clinic, Paracelsus Medical University, Salzburg, Austria; Kathmandu University School of Medical Sciences, Dhulikhel, Nepal; Research and Innovation Management (RIM) Department, Biostatistics and Publication of Clinical Trials, Paracelsus Medical University, Salzburg, Austria; Department of Ophthalmology and Optometry, Paracelsus Medical University, Salzburg, Austria; Research Program Experimental Ophthalmology and Glaucoma Research, Paracelsus Medical University, Salzburg, Austria; Center for Public Health Care Research, Paracelsus Medical University, Salzburg, Austria

**Author notes:** Correspondence to Kathrin Bogensberger.

**Keywords:** osteoarthritis, depression, risk factors, epidemiology, comorbidity

## Abstract

**Introduction:** Globally, osteoarthritis (OA) is the most prevalent musculoskeletal disease, affecting approximately 600 million people. It is characterised by progressive degeneration of the articular cartilage, osteophyte formation, and asymmetric joint space narrowing, leading to pain, stiffness, and functional impairment. The current focus of disease management is on symptom relief and functional improvement. However, these interventions frequently do not provide adequate outcomes. This may be attributed to a lack of consideration for contextual factors, including the presence of comorbidities such as depression. Several studies indicate that depression is highly prevalent, affecting up to 20% of OA patients. As depression is known to be a multifactorial disorder, there are various risk factors that may increase the likelihood of comorbid depression in patients with OA. Consequently, the scoping review’s objective is to map the existing literature on risk factors for comorbid depression in individuals with OA. Furthermore, the review is expected to provide important information for further in-depth investigation and the development of predictive tools to support the early identification of patients with OA at risk of comorbid depression.

**Methods and Analysis:** The intended review will target studies reporting risk factors for comorbid depression in individuals with OA. All available primary and grey literature will be considered for inclusion. Only articles published in English or German will be included in the review. The review will follow the format specified by the Preferred Reporting Items for Systematic Reviews and Meta-Analyses Extension for Scoping Reviews (PRISMA-ScR) and the JBI Manual for Evidence Synthesis. Original research in published and unpublished literature from inception until August 2024 will be included into the review. The databases to be searched will include PubMed, EMBASE, PsychInfo, and Web of Science. In addition, further literature will be identified by searching the reference lists of the included studies. Three reviewers will independently screen the identified studies for final inclusion. The data will be extracted and presented in tabular form and in a narrative summary that aligns with the review’s objective. Furthermore, a quality assessment of the included studies will be conducted using appropriate tools, and the results will be incorporated into the synthesis. Any disagreements that arise between the reviewers will be resolved through discussion or with an additional reviewer.

**Ethics and Dissemination:** Ethics committee approval will not be required because only published and publicly available data will be examined to answer the research question of the scoping review. We will not collect any personal information or any information that requires ethical approval.

**Strengths and Limitations:**

- This protocol adheres to the Preferred Reporting Items for Systematic Reviews and Meta-Analyses extension for Scoping Reviews checklist format, thereby ensuring best practice in conducting this review.
- The broad eligibility criteria and the conduct of the search in multiple databases with publication dates starting at inception will provide a comprehensive overview on the research area.
- The review will be conducted by a multidisciplinary team.
- A quality assessment of the included studies will facilitate a more accurate interpretation of the review’s findings and their significance for policy and practice.
- The review is limited to articles published in English and German.

## Introduction

Osteoarthritis (OA) represents the most prevalent musculoskeletal disease, with an estimated prevalence of approximately 600 million people worldwide (1). It is characterised by a progressive degeneration of the articular cartilage, osteophyte formation, and asymmetric joint space narrowing, which result in significant pain, stiffness, and functional impairment (1, 2). With the increase in life expectancy, OA is predicted to become more prevalent in the coming decades (3). People are experiencing more years with disability and are perceiving adverse impacts on their health and wellbeing. Chronic diseases, such as OA, with its disabling conditions and the associated increase in pain can impair activities of daily living and affect patient’s quality of life (2). OA is also known to negatively impact (work-) participation and increase healthcare utilisation, making it a significant economic and individual burden that should be considered from a public health perspective (1). Current management strategies aim to reduce symptoms and improve function (4). However, many of the more generalised interventions, do not provide sufficient symptom relief on an individual level. One explanation for such differences in effectiveness could be the presence of comorbidities such as depressive disorders (2, 4). It is evident that people with OA are at greater risk of experiencing mental health issues (5). Several studies indicate that depression is highly prevalent among patients with OA, affecting up to 20% of them (6). In comparison, the prevalence of depression in the European general population ranges from 3-10%, with Austria having a prevalence of 4% (7, 8). The coexistence of OA and depressive disorders can have a significant impact on individuals, the economy, and society (9). Depressive illness may hinder treatment approaches, such as physical activity, which is recommended to reduce pain intensity and disability in patients with OA. This can result in increased pain perception and less successful outcomes (2). Furthermore, those who are affected are at a higher risk of increased healthcare utilisation and reduced treatment satisfaction (5, 6). Although mental health is widely recognised as an important factor in managing patients with OA, a majority of those affected do not receive any mental healthcare support when suffering from co-occurring depressive disorder (2). Undertreatment of this co-occurring conditions has been shown to ignit a vicious cycle of increased pain intensity and the depressive illness. The interaction of these two chronic conditions has a negative impact on the course of each disease and the patient’s pre-existing OA severity, affecting their quality of life, the perception of the disease and its outcomes (10).

In order to prevent comorbid depression in individuals with OA and the widespread consequences of the co-occurrence of these diseases, optimal treatment and early detection is crucial. As depression is known to be a complex multifactorial disorder, various risk factors have been described to distinguish between OA patients with and without depressive symptoms. In accordance with a narrative review by Wang and Ni (6), the association between OA and depression is not unidirectional causal but rather bidirectional with each of the two diseases being an invigorating factor for the disease and symptom state of the other. The review identified fatigue, social contact, physical impairment, and BMI as predictors, as well as female sex, experiencing higher levels of pain, and stressful life events (6). These findings were further supported by a recent systematic review of 121 studies conducted by Fonseca-Rodrigues et al. (10), which highlighted the association between OA symptom burden, particularly pain and functional disability, and the development of depression. Furthermore, additional evidence has indicated that an elevated number of comorbidities, low income, and smoking also emerge as strong contributors to an increased risk of depression (9, 11). Additionally, there is evidence of specific comorbidity pathways, indicating that numerous predictors contribute to the development of depression in individuals with OA, thereby demonstrating a biological link between these chronic conditions (6, 12). These findings are in accordance with those of Zhang et al. (13), who provided further evidence for the existence of biomarkers. Consequently, serum brain-derived neurotrophic factor (BDNF), vascular endothelial growth factor (VEGF), and S100B might serve as potential biomarkers for identifying major depressive disorder (MDD) in patients with OA. Furthermore, serum BDNF and S100B have demonstrated considerable potential in predicting the risk of MDD in OA patients.

### Objective and Review Question

The primary objective of this scoping review is to provide a comprehensive identification and categorisation of risk factors associated with comorbid depression in individuals suffering from OA. It is planned to focus on biological, psychological, socio-psychological, and social risk factors. Furthermore, the review will map the identified factors and their relationship in the context of comorbid depression in OA. In the process of developing our scoping review protocol, we identified a systematic review focussing on the topic of “Risk Factors for Depression in Patients with Knee Osteoarthritis. A Systematic Review and Meta-Analysis” that had already been registered in PROSPERO (ID: CRD42023425840). The objective of that systematic review is to identify the risk factors for comorbidity depression in knee OA patients. While systematic reviews are typically focused on synthesising and critically appraising the evidence to answer specific research questions, the purpose of the scoping review presented in the current manuscript is broader in scope. The scoping review will encompass a wider range of study designs, including not only cohort and case-control studies but also other relevant study types, such as cross-sectional studies. Furthermore, the search will be extended to include APA PsychInfo, which is not covered in the systematic review. This will ensure a more comprehensive capture of the literature. The broader inclusion criteria and extensive search strategy will enable us to map the available literature more thoroughly and provide an overview of the extent and nature of research in this field. The chosen methodology serves to complement the systematic review by providing a comprehensive examination of the entire literature. This may potentially uncover areas that may benefit from further in-depth investigation and provide guidance for future systematic reviews. The following research question has been formulated: *“What are known biopsychosocial risk factors for comorbid depression in individuals with osteoarthritis?”* The results of the scoping review will provide comprehensive information about the risk factors related to the onset of comorbid depression in people with OA.

## Methods and Analysis

The scoping review protocol has been pre-registered in the Open Science Framework database (Registration DOI: https://doi.org/10.17605/OSF.IO/RYJH6) in compliance with the reporting guidance provided in the Preferred Reporting Items for Systematic Reviews and Meta-Analyses Extension for Scoping Reviews (PRISMA-P) (14). The proposed scoping review will be reported in accordance with the reporting guidance provided in the Preferred Reporting Items for Systematic Reviews and Meta-analyses extension for Scoping Reviews (PRISMA-ScR) (15). The research objectives, inclusion criteria, and methodological techniques will be determined prior to study commencement using the Joanna Briggs Institute Reviewers’ Manual 2020 Methodology for JBI Scoping Reviews (16). The process will adhere to the indicated framework, which comprises the following steps: [1] identifying the research question (see above); [2] identifying relevant studies; [3] selecting studies; [4] charting data, and [5] collating, summarising and reporting the results. The team of reviewers conducting the scoping review will be multidisciplinary, comprising a range of different disciplines. Studies of poor quality are often prone to a high risk of bias. Consequently, the findings of such studies are frequently questioned by researchers, practitioners and policymakers. It is therefore of the utmost importance to critically evaluate every included study in order to assess the risk of bias and the reliability of the results. On this line and in accordance with the recommendations set out by Levac et al.(17), a quality assessment will be included as an additional step of the scoping review.

## Inclusion Criteria

This scoping review pertains to the PEO framework with the objective of identifying the risk factors associated with comorbid depression in individuals with osteoarthritis.

### Participants (P)

This review will include available literature on participants aged 18 years or above, suffering from all forms of OA.

### Exposure (E)

The aim of the review will be to identify risk factors for the onset of comorbid depression among individuals with osteoarthritis. These risk factors will be classified as characteristics, conditions, behaviours, and other factors that increase the likelihood of comorbid depression as the outcome. The demographic factors to be explored include characteristics such as age, sex at birth, and ethnicity. Behavioural-related factors may include, but are not limited to, the level of physical activity, the consumption of tobacco products, and alcohol. In addition, socio-economic factors such as education level, employment status and social status are also taken into account in the review. Social factors also include the broader socio-cultural and environmental context and factors related to the social environment, such as social support in the personal environment and marital status. Furthermore, comorbidities, medications, clinical factors, and factors related to disease severity of OA (e.g. pain, functional factors) will be included in the review scope. Biological factors will include physiological and genetic aspects as well as biomarkers related to comorbid depression in OA. Sociopsychological and psychological factors refer to individual characteristics, which may include a history of trauma, substance abuse, personality disorders, mental health issues, social isolation, and stressful life events.

### Outcome (O)

The outcome of interest will be comorbid depression in individuals with osteoarthritis. Studies evaluating depression within the context of bipolar disorder, the perinatal period, schizoaffective disorder and premenstrual dysphoric disorder will be excluded.

#### Types of Data Sources

This scoping review will consider primary studies published from inception until August 2024. Quantitative and qualitative studies, whether published or unpublished, will be considered for inclusion. Quantitative studies encompass a range of experimental and quasi-experimental study designs, including randomised controlled trials, non-randomised controlled trials, before and after studies, and interrupted time-series studies. Furthermore, analytical observational studies, including descriptive studies, cohort studies, case-control studies, analytical cross-sectional studies and longitudinal studies will be considered for inclusion. In addition, grey literature will be sought, for example, on preprint servers (e.g. EuropePMC, PsyArXiv, MedRxiv, JMIR Preprints), Google Scholar, and trial registers. Only English and German language studies will be considered for inclusion.

#### Exclusion Criteria

Commentaries, correspondences, editorials, books and book chapters, lectures, and animal studies will be excluded. It should be noted that all forms of reviews will be excluded from the study. However, the reference lists will be examined in order to identify any additional articles that may meet the objectives of the reviews. There will be no limitations regarding risk factors for depression in individuals with OA in different geographical locations or specific racial or gender groups.

### Information Sources and Search Strategy

The search strategy was developed from the first author (KB) and was peer reviewed by the additional authors. As mentioned above, the aim of the scoping review is to generate a better understanding of the current research landscape on risk factors for comorbid depression in people with OA. The search strategy will aim to locate published and unpublished primary studies that fit within the eligibility criteria. An initial limited search of PubMed was undertaken to identify evidence on the topic. The text words contained in the titles and abstracts of relevant papers were used to develop a full search strategy for the scoping review. Alternative terms and combinations were identified by analysing keywords from preceding articles relevant to this review with the text mining web based reading and analysis environment for digital texts Voyant©. The strategy includes all identified keywords and index terms and will be translated to EMBASE, Web of Science, and PsychInfo. The references of the included articles will be hand searched to identify any additional evidence sources. The scoping review will include all studies published in English or German. In addition, grey literature will be searched using the same keywords. For further details on the PubMed search strategy, please refer to table 1.

**Table 1:** PubMed Search Strategy

| Bundle | Keywords searched |
| --- | --- |
| Population (Individuals with OA) | Osteoarthritis [mh]<br>Osteoarthritis [tiab]<br>Osteoarthros* [tiab]<br>Degenerative arthr* [tiab]<br>or/1-4 |
| Risk factor(s) for comorbid depression | Risk [mh]<br>Epidemiology [mh subheading]<br>Epidemiologic factors [mh]<br>Epidemiological studies [mh]<br>Etiology [mh subheading]<br>correlat* [tiab]<br>determinant* [tiab]<br>risk factor* [tiab]<br>prognos* [tiab]<br>relationship* [tiab]<br>prediction [tiab]<br>predict* [tiab]<br>association* [tiab]<br>prevalence [tiab]<br>or/6-19 |
| Outcome (depression) | Depression [mh]<br>Major depressive disorder [mh]<br>Depression [tiab]<br>Depressive (disorder OR symptom*) [tiab]<br>or/21-24 |
|  | 5 and 20 and 25 |

*mh* (MeSH term, subject heading in PubMed); *tiab* (title, abstract search in PubMed), *mh subheading* (MeSH term subheading in PubMed)

### Screening

Following the search (KB), EndNote version 20 will be used for reference management and deduplication. Rayyan© (URL: https://www.rayyan.ai/) will be utilised to organise, manage, and accelerate the scoping review screening process. Three reviewers (KB, DSS, EP) will independently conduct the title and abstract screening for assessment of the inclusion criteria for the review. If the reviewers have uncertainties about the relevance of a study or if the abstract is unclear, the full article will be retrieved. Subsequently, the full texts will be examined in detail and screened for inclusion by the same three reviewers. Third, references of all considered articles will be hand searched to identify any relevant report missing in the search strategy. Any disagreements will be resolved by discussion or the inclusion of an additional reviewer (AvZ). The results of the search and the study inclusion process will be reported in full in the final scoping review and presented in a Preferred Reporting Items for Systematic Reviews and Meta-analyses extension for scoping review (PRISMA-ScR) flow diagram (15).

### Data Extraction

The extraction of relevant data from the included studies, will be aided by the use of an organised charting process developed a priori (KB). A table of the proposed data extraction form is shown in table 2. The form will be charted on an Excel spreadsheet. The data extraction form will be piloted by three independent reviewers (KB, DSS, EP) using a random sample of 10% of the eligible articles to ensure consistency and accuracy. Clarification and updating of the extraction form will be an iterative process until all authors reach consensus on the final version. If necessary, the data extraction tool will be continually reviewed as part of this process. Data extraction will be undertaken independently by the three reviewers. If appropriate, authors of papers will be contacted to request missing or additional data, where required. Descriptive summary tables will be produced to recapitulate the evidence base.

**Table 2:**
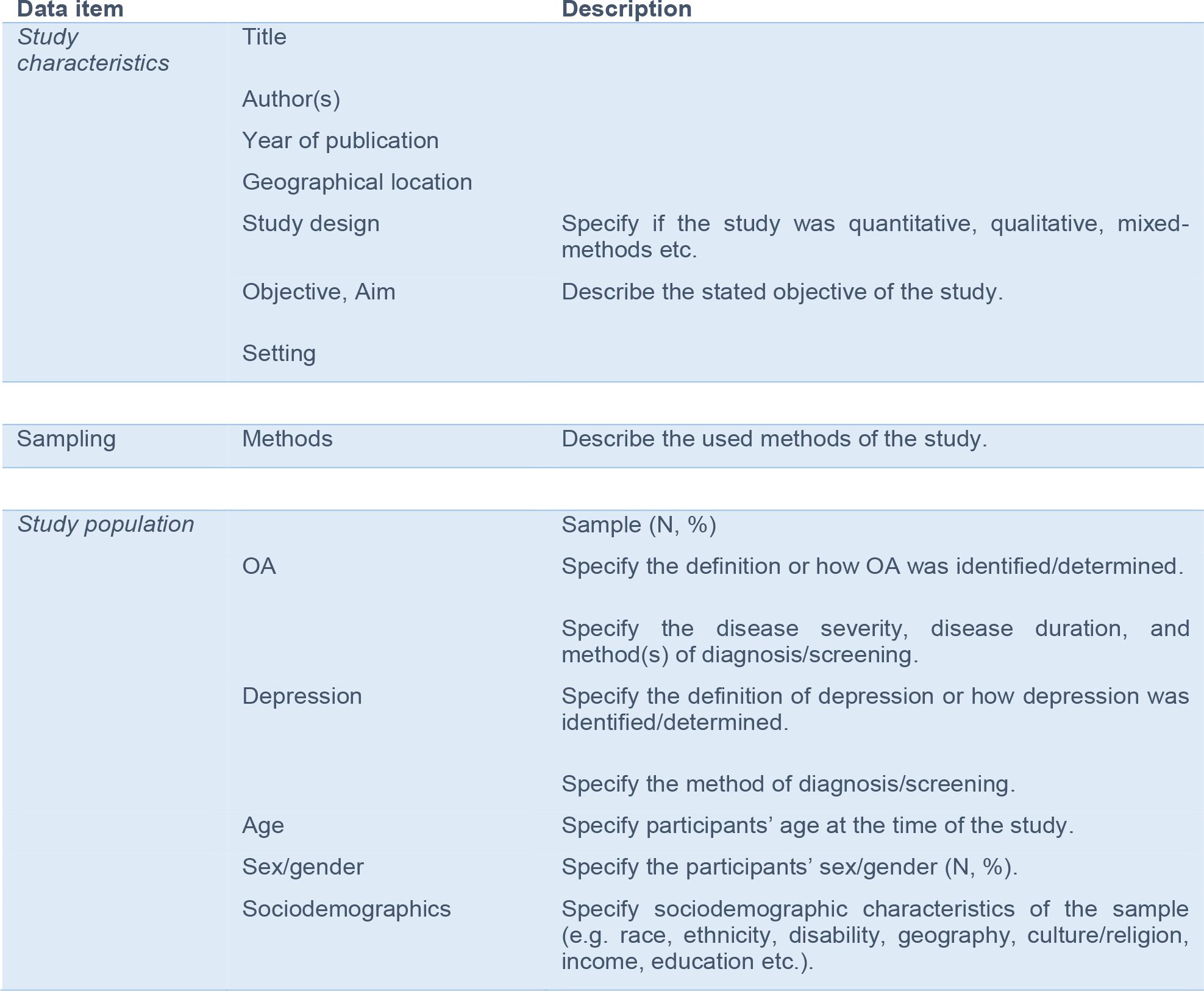
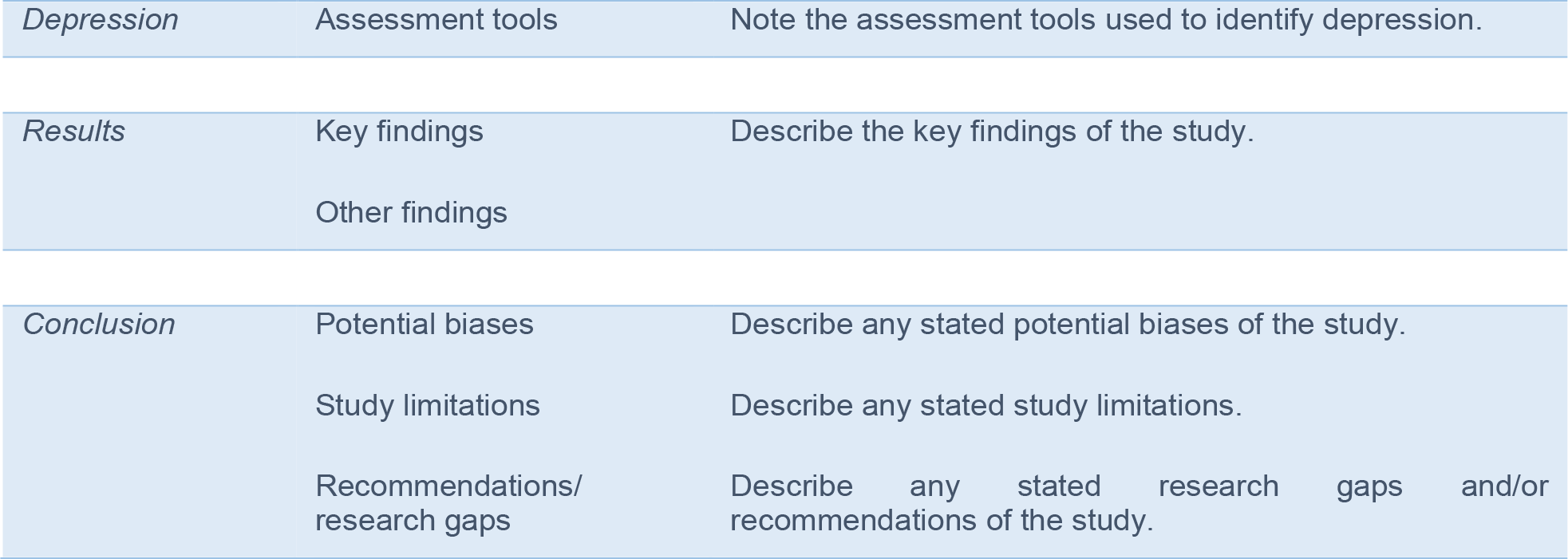
Charting table

### Risk of Bias Assessment

As Levac, Colquhoun & O’Brien (17) have already highlighted, the necessity for quality assessment in a scoping review should be addressed, as otherwise the findings of the review are difficult to interpret and limits arise for the uptake of the scoping review findings into policy and practice. In order to enhance the rigor and reliability of this scoping review, a quality assessment of the included studies will be conducted. The Cochrane Risk of Bias Tool will be used for randomised controlled trials, the STROBE checklist for observational studies, the MINORS checklist for quasi-experimental studies, the Critical Appraisal Programme Checklist for qualitative studies, and the Mixed Methods Appraisal Tool for mixed-methods studies. In addition, the AACODS checklist will be used to evaluate diverse formats of grey literature. The quality of each included study will be independently assessed by three reviewers (KB, DSS, EP) using the selected tools. Any discrepancies will be resolved through discussion and in case of sustained disagreement through the involvement of an additional reviewer (AvZ). The quality assessment data will be extracted using a standardised form. The results of the quality assessment will be used to provide a narrative summary of the quality of the evidence. Although studies will not be excluded based on the results of the quality appraisal, the quality will be taken into account when interpreting the findings.

### Data Synthesis

The outcomes and other information collected regarding the selected studies will be synthesised using both quantitative and qualitative methods. This will be accompanied by a narrative summary of the findings. The synthesis will be presented in tabular form, with summary data presented graphically, and individual data for each study presented in tabular form. The overarching objective of the synthesis is to identify the risk factors that have been identified in the current body of knowledge. Furthermore, the objective of the scoping review is to identify research gaps and present recommendations for future research agendas.

## Conclusion

The purpose of this scoping review is to map the existing literature on the risk factors influencing the comorbidity of depression and OA. Understanding these factors will help to identify deficiencies in patient care and research. The findings of this review will also provide valuable insights into the areas where further action is required to support patients with OA at high risk of experiencing comorbid depression. Moreover, the scoping review may assist researchers in developing and confirming their preliminary inclusion criteria and the research questions for subsequent systematic reviews and meta-analyses by identifying available and relevant evidence. Furthermore, the scoping review may support the development of predictive tools to facilitate the early identification of patients with OA at risk of comorbid depression, and may also unravel pathophysiological conditions functionally linking OA and depression.

## Data Availability

Further details regarding the present scoping review protocol can be obtained by contacting the authors.

## Ethics and Dissemination

Ethics committee approval will not be required because only published and publicly available data will be examined to answer the research question of the scoping review. We will not collect any personal information or any information that requires ethical approval.

## Support and Funding

This research did not receive any specific grant from funding agencies in the public, commercial, or not-for-profit sectors.

## Conflicts of Interest

The authors declare that they have no competing interests.

## Acknowledgements

We thank the reviewers and editors for their assistance. Furthermore, we would like to express our gratitude to Tim Johansson, Anna Winkler, Roland Essl-Maurer, and Cvetka Lipuš for their professional support.

## Author’s Contributions

KB and AvZ conceptualised the project. KB will be acquiring, analysing and interpreting the data. DSS and EP assisted in the refinement of the conceptualisation of the project, particularly with regard to the methods section. They will also be involved in acquiring the data and interpreting the data. KB drafted the manuscript. All authors revised the manuscript critically and agreed to the commencement of the scoping review and the submission of the current protocol. All authors agree to be accountable for all aspects of the work, ensuring that any questions related to the accuracy or integrity of any part of the work are appropriately investigated and resolved. All authors will be involved in the final approval of the published version.

## Notes

### Competing Interest Statement

The authors have declared no competing interest.

### Funding Statement

This study did not receive any funding.

